# A phase I, open-label, multi-center dose-finding and expansion study to investigate the safety, tolerability, and preliminary efficacy of CR-001 (5-FU-miR-15a) in patients with acute myeloid leukemia

**DOI:** 10.1101/2025.06.08.25328885

**Authors:** Irini Ktoridou-Valen, Andrea Lenartova, Sturla Magnus Grøndal, Cara E. Wogsland, Adam S. Dowrick, Mark Wood, Knut T. Smerud, Nils Meland, Thomas Dahl, Andrew Fesler, Jingfang Ju, Bjørn T. Gjertsen

**Affiliations:** K.G. Jebsen Centre for Myeloid Blood Cancer CMYC, Department of Clinical Science, University of Bergen, Bergen, Norway; Center for Cancer Biomarkers CCBIO, Department of Clinical Science, University of Bergen, Norway; Haukeland University Hospital, Department of Medicine, Hematology section, Norway; Oslo University Hospital, Department of Hematology, Norway; KinN Therapeutics AS, Norway; Smerud Medical Research International AS, Norway; Curamir Therapeutics, Inc.; Stony Brook University, Department of Pathology, Renaissance School of Medicine

## Abstract

**Purpose:** Evaluating the safety and tolerability of 5-FU-miR-15a (CR-001) in adults with relapsed or refractory acute myeloid leukemia (R/R AML) in a phase I dose-escalation study (EudraCT number: 2021-006332-46).

**Patients and Methods:** A standard 3+3 dose-escalation study design was employed. Patients received intravenous (IV) administrations of the synthetic double-stranded mimic of miR-15a (miRNA) CR-001 of 7.5 mg, 11.25 mg, 15 mg and 18.75 mg/administration, weekly dosing. Each patient was dosed on a single dose level. The formulation included a fixed dose of GMP-grade linear polyethyleneimine (PEI; α-Methyl-ω-hydroxy-poly[(iminioethylene)chloride]; Poly[imino(1,2-ethanediyl)]; CAS Number 26913-06-4; EC Number 608-018-6) as the excipient: 15 mg of PEI with 7.5 mg CR-001, and 20 mg of PEI with all higher dose levels. Glucose was added to stabilize the miRNA/PEI complexes. Biomarker analysis was performed using mass cytometry (CyTOF) on peripheral blood and bone marrow samples to quantify surface and intracellular protein expression at the single-cell level.

**Results:** A total of 11 patients with R/R AML (median age: 74 years; all classified as adverse risk per ELN 2022) received a cumulative 66 intravenous doses of CR-001 (range: 1–16 doses; median: 5) across the four dose levels. The treatment was well-tolerated. One patient demonstrated a partial clinical response, with resolution of extramedullary pericardial AML after four doses. Among the 10 evaluable patients, six achieved stable disease (SD) according to ELN criteria after four infusions. Single-cell profiling mass cytometry revealed molecular activity of CR-001 on known targets of miR-15a (e.g., BMI1, WEE1 and MCL-1) beginning at the lowest dose level (7.5 mg). Concurrently, an increase in BAX, a pro-apoptotic marker, was observed following treatment.

**Conclusions:** CR-001 was well tolerated and demonstrated biological activity as evidenced by target modulation and disease stabilization in R/R AML patients. Biological activity was observed starting at the lowest dose level (7.5 mg/administration), with initial signs of clinical efficacy at 11.25 mg per administration. This trial was closed prematurely due to funding constraints, however, based on these findings further development is warranted, possibly in therapy combinations.

## INTRODUCTION

Acute myeloid leukemia (AML) accounts for approximately 1% of all malignancies diagnosed in the United States, with an estimated incidence of over 20,000 new cases in 2024. ^1^ AML is a genetically and clinically heterogeneous disease characterized by a high rate of relapse and limited long-term survival in most patients. ^2^ Therapeutic resistance is one of the major reasons for the refractoriness and relapse of the disease. ^3^ Consequently, novel therapeutic options are urgently needed to improve patient outcomes and prolong AML patient survival by sustaining complete remission (CR). Notably, only 10% of relapsed or refractory disease patients survive beyond 5 years. ^4^

MicroRNAs (miRNAs) have been extensively investigated as key epigenetic regulators in both solid tumors and hematological malignancies. ^5^ miRNAs are short non-coding RNAs that regulate target gene expression post-transcriptionally through interaction with the 3’ untranslated region (3’UTR) of target mRNA. ^6,7^ This interaction results in either mRNA degradation or translational repression, thereby decreasing the expression of target proteins. A single miRNA can regulate multiple mRNAs. This makes miRNAs attractive therapeutic options, particularly for overcoming drug resistance, by enabling the simultaneous suppression of several genes involved in chemoresistance ^8^, despite challenges in delivery and stability that remain an active area of research. ^9^

Previous studies have shown that tumor suppressor miRNAs such as miR-15a play a pivotal role in hematological malignancies, including chronic lymphocytic leukemia (CLL), AML, and various solid tumor types by suppressing oncogenic targets and pathways including BCL-2, BMI-1, WEE-1, YAP1, CHK1, DCLK-1, and MCL-1. ^10–14^

We have developed a novel modification strategy to enhance the antitumor potential of tumor suppressor miRNAs through conjugation with chemotherapeutic agents, such as the cytotoxic pyrimidine nucleoside analogs 5-fluorouracil (5-FU) and gemcitabine. ^12,15,16^ Specifically, modification of miR-15a with 5-FU has resulted in the creation of the 5-FU-miR-15a (CR-001) that has demonstrated enhanced therapeutic efficacy, stability, and deliverability across multiple tumor types in preclinical models ^14^ (**Figure 1)**.

**Figure 1.**
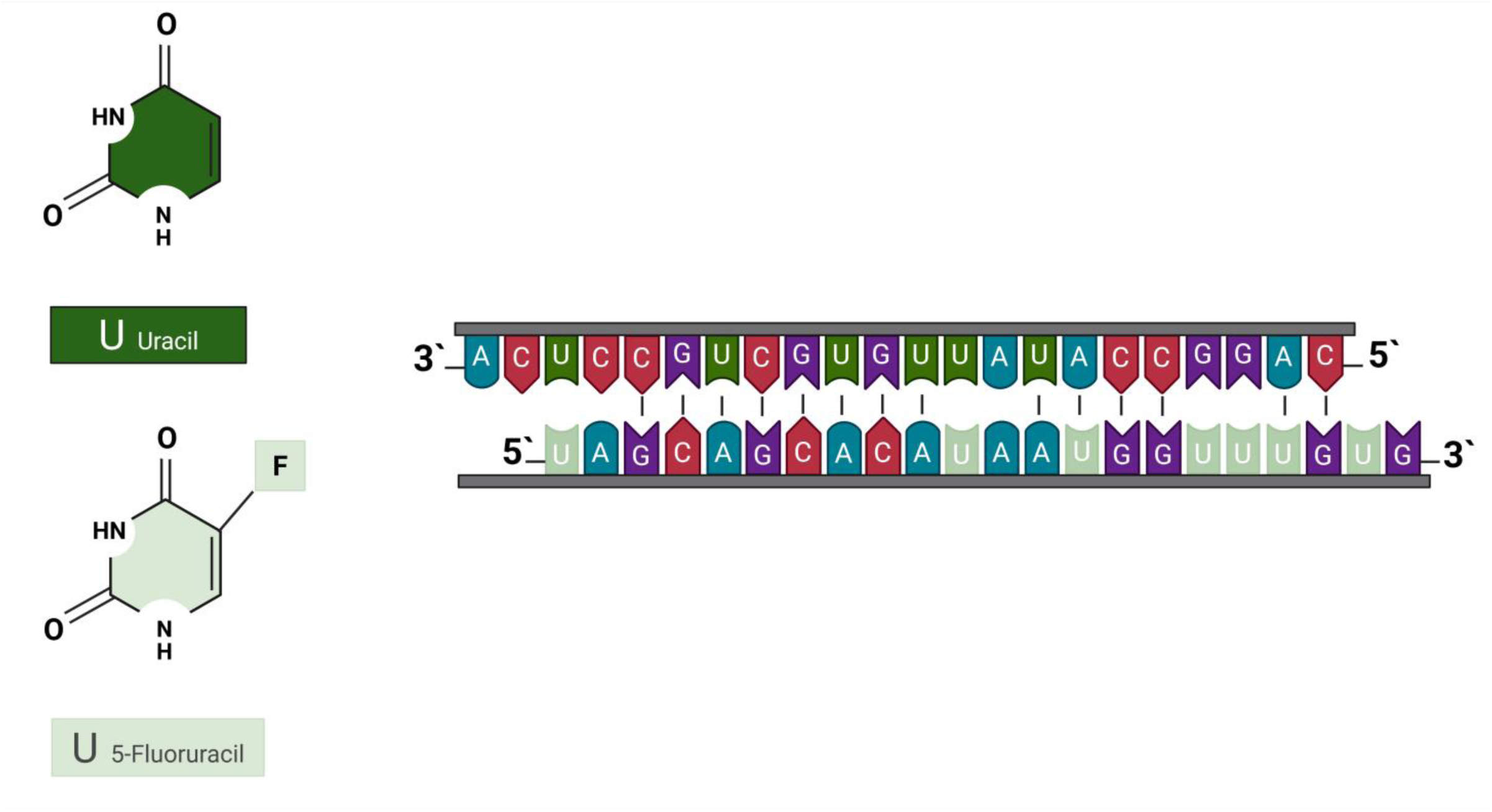
The sequence the tumor suppressor miRNA, miR-15a, with its uracil bases of the guide strand of the mature miRNA substituted with the antimetabolite <u>nucleoside analog</u> 5-fluorouracil (5-FU). Adapted from: John G. Yuen, Andrew Fesler, Ga-Ram Hwang, Lan-Bo Chen, Jingfang Ju, Development of 5-FU-modified tumor suppressor microRNAs as a platform for novel microRNA-based cancer therapeutics, Molecular Therapy, 2022. https://doi.org/10.1016/j.ymthe.2022.07.015.

Here, we report the first dose-finding and toxicity evaluation of the 5-FU-miR-15a CR-001 in humans, suggesting that miRNA used as intravenous therapeutic is tolerable in AML (EudraCT number: 2021-006332-46).

## METHODS

### Patients

Between 15 August 2022 and 13 December 2023, a total of 11 patients were enrolled and treated with a total of 66 doses of CR-001. The baseline characteristics of all participants are summarized in **Table 1**. There were 10 (91%) patients with relapsed/refractory AML and 1 (9%) patient with newly diagnosed AML considered as not being eligible for standard of care by Principal Investigator (PI).

**Table 1.** Baseline demographic and clinical characteristics of the patients. * Easter Cooperative Oncology Gmup (ECOG) perfonmance status scores range from 0 to 5, with 0 indicating no symptoms and higher soo11es indicating greater disability. * AML denotes acute myeloid leukemia, MDS myelodysplastic syndrome, MP1N myelapmlifemtive neoplasia. † ELN European Leukemia Network.

| CR-001 dose (mg) | 7.5 | 11.25 | 15 | 18.75 | All |
| --- | --- | --- | --- | --- | --- |
| No. of patients in each dose level | N=4 | N=3 | N=3 | N=1 | N=11 |
| **Characteristic** |  |  |  |  |  |
| Age median | 74 | 61 | 74 | ~70 | 74 |
| Male gender - n (%) | 2 (50) | 2 (66) | 1 (33) | 0 (0) | 5 (46) |
| *ECOG (0-1) - n (%) | 2 (50) | 2 (66) | 3 (100) | 1 (100) | 6 (55) |
| Bone marrow aspirate blast count at baseline - median (range) | 20 (17-45) | 25 (9-43) | 16 (15-16) | 49 | 20 (9-45) |
| **Disease characteristics at last assessment prior to study start** |  |  |  |  |  |
| Adverse genetic risk by †ELN2022 - n (%) | 4 (100) | 3 (100) | 2 (66) | 0 (0) | 9 (82) |
| TP53 mutation - n (%) | 2 (50) | 2 (66) | 1 (33) | 0 (0) | 5 (46) |
| Complex karyotype - n (%) | 2 (50) | 2 (66) | 2 (66) | 0 (0) | 6 (55) |
| **AML secondary to MPN/MDS - n (%) | 4 (100) | 2 (66) | 1 (33) | 0 (0) | 7 (64) |
| Therapy related AML - n (%) | 0 (0) | 1 (33) | 1 (33) | 0 (0) | 2 (19) |
| **Disease status assessment at study start - n (%)** |  |  |  |  |  |
| Untreated | 1 (25) | 0 (0) | 0 (0) | 0 (0) | 1 (9) |
| First relapse | 1 (25) | 0 (0) | 0 (0) | 1 (100) | 2 (19) |
| Second relapse | 0 (0) | 0 (0) | 1 (33) | 0 (0) | 1 (9) |
| Third relapse | 3 (75) | 3 (100) | 2 (66) | 0 (0) | 8 (73) |
| Extramedullary disease | 0 (0) | 1 (33) | 1 (33) | 0 (0) | 2 (19) |
| Median number of previous systemic therapies - n (range) | 2 (0-3) | 3 (3-4) | 3 (2-3) | 1 | 3 (0-4) |
| Relapsed post intensive chemotherapy - n (%) | 2 (50) | 3 (100) | 2 (66) | 0 (0) | 7 (64) |
| Relapsed post allogeneic stem cell transplant - n (%) | 2 (50) | 3 (100) | 2 (66) | 0 (0) | 7 (64) |
| Relapsed post second allogeneic stem cell transplant - n (%) | 0 (0) | 1 (33) | 0 (0) | 0 (0) | 1 (9) |
| Refractory/relapsed post venetoclax + azacitidine - n (%) | 3 (75) | 2 (66) | 2 (66) | 1 (100) | 8 (73) |

All patients had an adverse risk genetic profile (ELN 2022) ^17^, 5 (43%) patients had a *TP53* mutation, and 6 (55%) patients had a complex karyotype.

The median age was 74 years (range 60-86). The study population was heavily pretreated with a median of three lines of therapy (range 0-4), and 7 (64%) patients had relapsed after allogeneic hematopoietic stem cell transplant (allo-HSCT). One patient, who relapsed after a second allo-HSCT, received eight doses of CR-001 as part of the study and an additional eight doses under compassionate use. All patients who received at least one dose of CR-001 were included in the safety analysis, whereas only patients who received at least three doses were included in the efficacy analysis.

### Study design and drug administration

This first-in-human, open-label, phase I, dose-escalation study was conducted at two sites in Norway and enrolled patients with R/R AML who were either ineligible for or had exhausted standard therapeutic options.

The investigational agent, CR-001, is a synthetic, double-stranded microRNA-15a (miR-15a) mimic chemically modified with 5-fluoruracil (miRNA-5FU) and formulated with the excipient PEI in glucose solution for stabilization of the miRNA/PEI complexes.

The primary objectives of the study were to assess the safety and tolerability of CR-001 and to estimate the maximum tolerated dose (MTD) or the recommended phase II dose (RP2D) of CR-001.

A standard 3+3 dose-escalation study design was employed. **Figure 2** illustrates the study design. The data presented here originate from a larger study that was terminated prematurely due to withdrawal of funding. In the original study design, CR-001 was to be evaluated across five dose levels during Part I (Dose Escalation), with level 5 corresponding to a dose of 22.5 mg.

**Figure 2.**
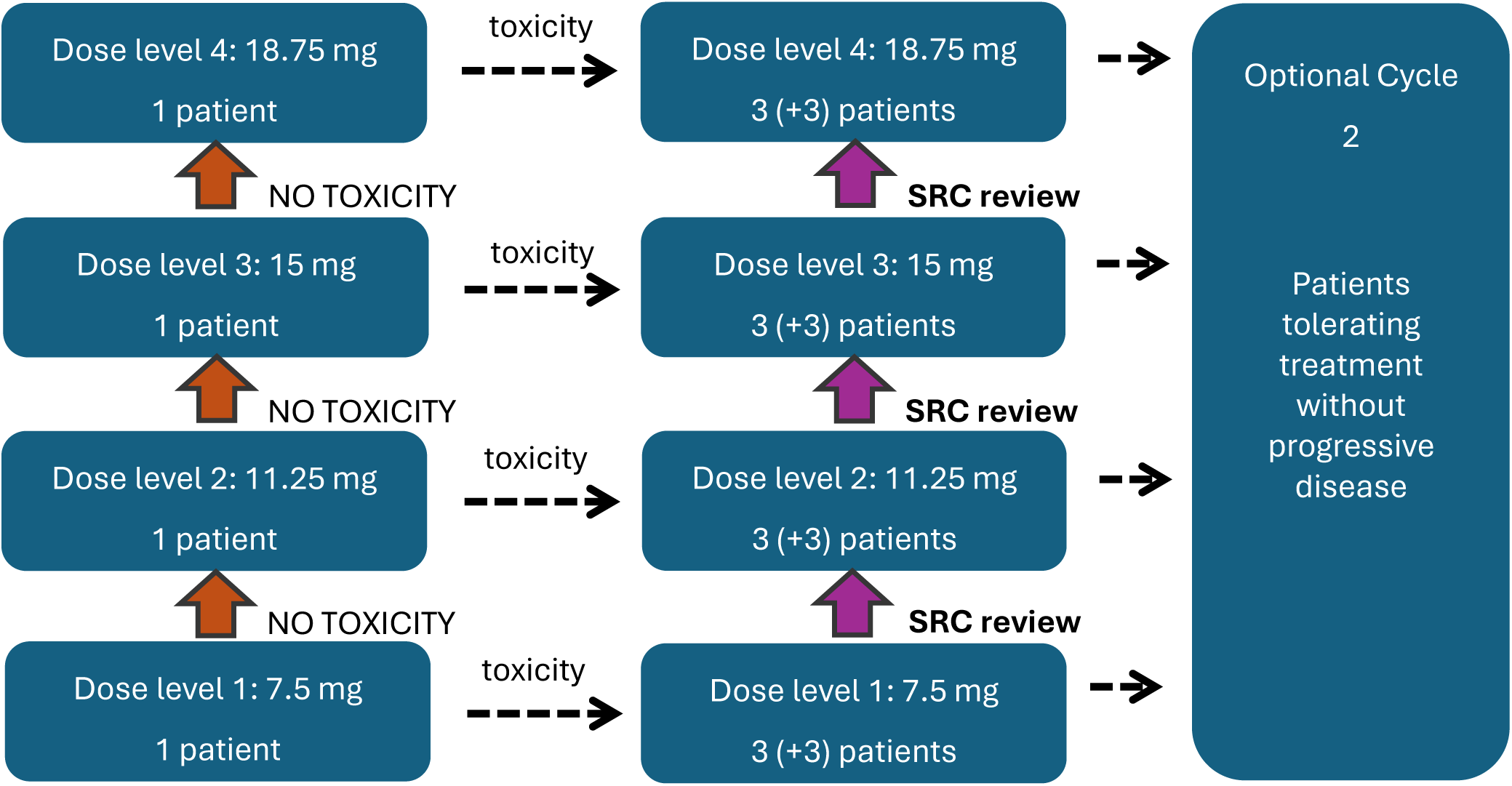
Study design, 3+3. Part I: Dose escalation. SRC; Safety Review Committee

However, only four dose levels were administered (only 1 patient on dose level 4) before study termination. The study was originally intended to proceed to Part II (Dose Expansion) following completion of the escalation phase.

Patients received intravenous (IV) administrations of CR-001 once weekly for four consecutive weeks at dose levels of 7.5 mg, 11.25 mg, 15 mg, or 18.75 mg per administration. PEI was co-administered at a fixed dose of 15 mg with the 7.5 mg CR-001 dose, and 20 mg with all higher dose levels (11.25 mg, 15 mg, 18.75 mg).

Patients received premedication with oral dexamethasone 20 mg administered 12–16 hours and again 1 hour prior to infusion, as well as paracetamol 1 g, famotidine 80 mg, and cetirizine 10 mg one hour before infusion to mitigate infusion-related reactions.

The efficacy evaluation was performed one week after the last administered dose, and safety was evaluated 30 days post-treatment. Patients who demonstrated stable disease after the initial four weekly doses were eligible to receive up to four additional doses of CR-001.

### Mass cytometry sample collection

Blood and bone marrow samples for mass cytometry analysis were collected at set time points with some variation (**Figure 3a**). Blood was collected at screening, before and 4 hours after the first dose, before dose on the remaining three weeks of cycle 1, at cycle 1 follow-up, and at safety follow-up (SFU) 30 days after the last dose. Bone marrow was collected at screening, on Day 29 (for cycle 1 follow-up), and at the start and follow-up of cycle 2.

**Figure 3a.**
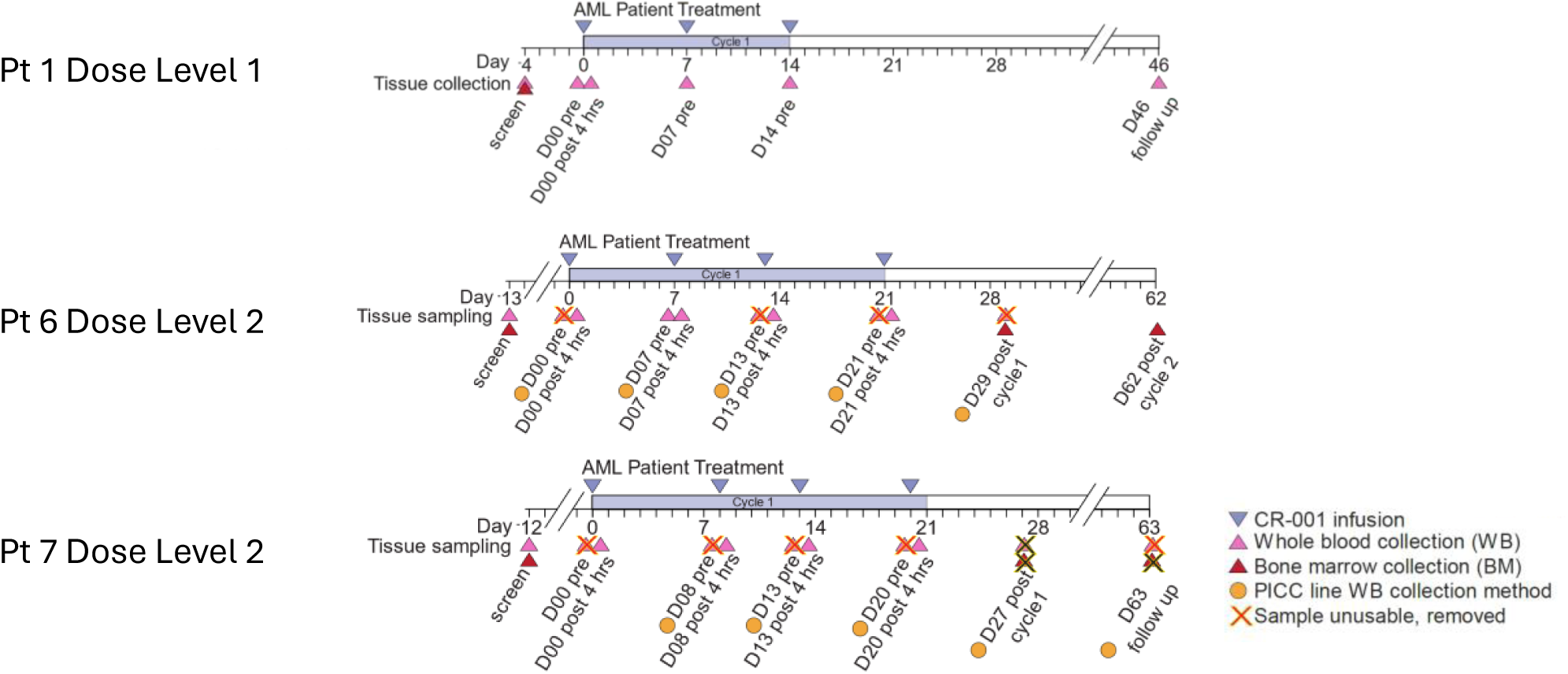
Sample collection timeline for patient 1, 6 and 7. Dose Level 1: 7.5 mg/administration, Dose Level 2: 11.25 mg/administration.

### Mass cytometry sample processing

An AML mass cytometry panel was designed to analyze the samples from patients treated with CR-001, as part of a Phase I clinical trial. The AML focused panel was developed to include the key surface markers for AML diagnosis and classification, detection of measurable residual disease (MRD), and six CR-001 targets of interest. The panel also contained antibodies against surface markers to classify other leukocyte cell types as well as several functional intracellular antibodies. ^18^ Pre-conjugated, mass-tagged antibodies were purchased from Standard Biotools (via AH diagnostics) where available. The antibodies: CD3, CD8, CD117, BAX, BCL-2, BMI1, MCL-1, and YAP were purchased in purified carrier-free form from other vendors and conjugated in-house to their respective mass tags using conjugation kits from Standard Biotools (**supplementary table 1**).

Samples were fixed using PROT1 (Smart Tube, Inc., Las Vegas, USA) according to manufacturer instructions and shipped on dry ice. Samples were thawed and processed according to manufacturer instructions. Samples were barcoded using Cell-ID 20-Plex Pd Barcoding Kit (Cat. # 201060, Standard Biotools) according to manufacturer instructions.

Control samples were prepared for batch anchor controls to be used in each stained barcoding batch and for antibody testing and titration. These controls included the validation anchor and the positive anchor. The validation anchor comprised healthy human whole blood containing approximately 7 million leukocytes per mL with a 10% spike of AML blast cells. This validation anchor was designed to serve as a reference point for the analysis. The positive anchor was designed to yield a positive signal for all antibodies in the panel. It was composed of approximately 60% CD45+ leukocytes from healthy whole blood from the same donor, 15% MOLM-13 AML cell line cells, 12% AML blast cells, and 12% BJ endothelial cell line cells. Validation and positive anchors were treated with Prot1 protein stabilizer (Smart tube, cat# 501351691 PROT1-1L), aliquoted, and frozen at -80°C. Aliquots were utilized as needed, including one of each for every patient analysis. Antibodies were tested and titrated on the CyTOF XT mass cytometer to achieve optimal staining concentrations. ^19^

Barcoded samples were pooled and stained with a 28-marker surface staining cocktail before being washed and permeabilized in ice-cold methanol for intracellular staining using an 11-marker panel (**Supplementary table 1**). After washing, cells were refixed in PFA and incubated with an iridium intercalator (Cat. # 201192B, Standard Biotools). Cells were frozen and kept at -80°C until acquisition, at which point the cells were thawed, washed with cell staining buffer (Cat. # 201068, Standard Biotools) and CAS-plus (Cat. # 201244, Standard Biotools) and acquired at 800 000 cells/mL.

### Mass cytometry analysis

CyTOF suspension mass cytometry data processing and analysis was performed on multiple platforms. Data was collected on the CyTOF XT (Standard BioTools). A MATLAB standalone debarcoding tool ^20^ was used for each batch to debarcode the data and split it into debarcoded sample-specific files. Minimum separation and Mahalanobis distance values are denoted in the FCS file names. The R package Premessa was used inside RStudio to concatenate files as needed. Data was gated in multiple iterations in Cytobank. Dimensionality reduction using tSNE-CUDA, data visualization and graphing, and both FlowSOM and SPADE clustering were all performed in Cytobank.

## RESULTS

Treatment of 11 AML patients (median age 74 y, all in adverse risk stratification ELN 2022) with CR-001 was well-tolerated through 66 doses (range 1-16, median 5) administered across the 4 dose levels. **Table 1** presents the baseline patient characteristics. **Table 2** presents the number of CR-001 infusions administered in each dose level and the 3 most frequent adverse events.

**Table 2.** Number of CR-001 infusions per dose level administered and tolerability. * Easter Cooperative Oncology Group (ECOG) performance status scores range from O to 5, with O indicating no symptoms and higher scores indicating greater disability.

| CR-001 dose (mg) | 7.5 | 11.25 | 15 | 18.75 | All |
| --- | --- | --- | --- | --- | --- |
| No. of patients in each dose level - N | N=4 | N=4 | N=3 | N=1 | N=11 |
| No. of doses administered in each level - n | n=12 | n=30 | n=16 | n=8 | n=8 |
| **Tolerability (evaluatable 11 patients)** |  |  |  |  |  |
| Toxicity evaluatable - n (%) | 4 (100) | 3 (100) | 3 (100) | 1 (100) | 11 (100) |
| *ECOG 0 -1 after 4 CR-001 infusions - n (%) | 1 (75) | 3 (100) | 3 (100) | 1 (100) | 8 (73) |
| Any adverse event - n (%) | 4 (100) | 3 (100) | 3 (100) | 1 (100) | 11 (100) |
| Infusion related reactions/all infusions administered - n (%) | 1/12 (8) | 3/30 (10) | 7/16 (44) | 0/8 (0) | 11/66(17) |
| Hyperglycemia per patient - n (%) | 3 (75) | 2 (66) | 2 (66) | 0 (0) | 7 (64) |
| Infection - n (%) | 4 (75) | 4(33) | 3 (66) | 1 (100) | 11 (100) |

### Tolerability

All 11 patients were evaluated for tolerability. There were no dose-limiting toxicities observed. All treated patients had an adverse event during treatment with CR-001. Listing of treatment-related adverse events (TRAEs) of any grade is shown on **Table 3**. The most frequent adverse event, irrespective of a relationship to CR-001 was infection (11 patients, 100%), and hyperglycemia (7 patients, 64%). Infusion-related reaction occurred in 6 (55%) patients. The symptoms occurred suddenly at the end of infusion or within 30 minutes after the infusion start. Shivering and tachycardia appeared in all 6 patients experiencing this event, fever only once in one patient. In all cases, the symptoms persisted for around 30 minutes and resolved completely. None of these 6 patients experienced hypotension or hypoxia. These 6 patients received hydrocortisone and antihistamine intravenously. Patient 9 that experienced fever received additionally an interleukin-6 blocker, tocilizumab, as cytokine-release syndrome was suspected. This patient experienced similar symptoms, but without fever, persisting for 30 minutes after each of the 7 subsequent CR-001 doses and chills about 4 days post infusion. Only this patient experienced infusion reaction more than once. Assessments were performed by the hospital pharmacies on the infusion bags of those patients with suspected cytokine-release syndrome (CRS). All infusion sets were found sterile. All patients with infusion reaction resumed treatment on dosing schedule. The infusion reaction appeared at all dosing levels higher than 7.5 mg and was observed besides prolonged infusion administration time from 20 to 120 minutes. No further mitigating strategies were developed or evaluated as the study terminated due to financing shortage.

**Table 3.** Treatment-related adverse events (TRAEs) This table presents only the treatment-emergent adverse events (TEAE) that were possibly probably related to CR-001; i.e., treatment-related adverse events (TRAEs). Other TEAEs defined as unlikely related to the IMP are not included in this table. CTCAE Common Terminology Criteria for Adverse Events. TEAE period was between the first dose of CR-001 and 30 days after the last dose.

| Treatment-related adverse events (TRAEs) | Grade (CTCAE) | Incidence of TRAE (n) | # of patients experiencing TRAE (n) | CR-001 dose (mg) |  |  |  |
| --- | --- | --- | --- | --- | --- | --- | --- |
|  |  |  |  | 7.5 | 11.25 | 15 | 18.75 |
| Insomnia | 2 | 1 | 1 |  |  |  |  |
| Intention tremor | 1 | 1 | 1 |  |  |  |  |
| Serum creatinine increase | 1 | 1 | 1 |  |  |  |  |
| Alanine aminotransferase increase | 1 | 5 | 1 |  |  |  |  |
| Alanine aminotransferase increase | 2 | 2 | 1 |  |  |  |  |
| Alanine aminotransferase increase | 3 | 2 | 1 |  |  |  |  |
| Aspartate aminotransferase increase | 1 | 2 | 1 |  |  |  |  |
| Aspartate aminotransferase increase | 2 | 1 | 1 |  |  |  |  |
| Alkaline phosphatase increase | 1 | 3 | 1 |  |  |  |  |
| Gamma-glutamyl transferase increase | 1 | 1 | 1 |  |  |  |  |
| Infusion- related reaction | 2 | 3 | 3 |  |  |  |  |
| Infusion-related reaction | 1 | 7 | 1 |  |  |  |  |
| Infusion-related reaction | 2 | 1 | 1 |  |  |  |  |
| Chills | 1 | 1 | 1 |  |  |  |  |
| Four days IMP post-infusion chills | 1 | 7 | 1 |  |  |  |  |
| Three days IMP post-infusion confusion and reduced physical ability | 1 | 1 | 1 |  |  |  |  |
| Cytokine-release syndrome | 1 | 1 | 1 |  |  |  |  |
| Difficult venous access | 3 | 1 | 1 |  |  |  |  |

In general, CR-001 was well-tolerated and administered on an out-patient basis. Eight (73%) patients were in ECOG 0-1 performance status after 4 doses of CR-001, including 3 (27%) patients who improved their performance from ECOG 2. Hyperglycemia was common (7 pts, 64%) and was attributed to high dose dexamethasone premedication. None of the patients had hyperglycemia at follow-up safety visit 30 days after the last dose .

### Preliminary efficacy

One patient (*NRAS* VAF 48%, *ASXL1* 41%, *TP53* 23%) responded with elimination of extramedullary pericardial AML disease after 4 doses of CR-001 but progressed in bone marrow after a total of 16 doses of CR-001 (**Figure 4**). This patient initially received eight weekly doses as part of the study. After approval from the Norwegian Medicines Agency (NoMA), the patient was given an additional eight doses under compassionate use. Following this, evaluation showed disease progression, and the patient was subsequently treated with trametinib as part of another clinical trial. Six of 10 evaluable patients experienced ELN stable disease (SD) after 4 CR-001 infusions; two of these patients continued to experience stable disease after 8 infusions. Three of 6 patients with SD exhibited *TP53* mutation. Both patients who experienced SD for at least 8 weeks had complex karyotype (CK) and *TP53* mutation. Single-cell profiling using mass cytometry uncovered signs of molecular activity of CR-001 on known targets of miR-15a (e.g., BMI1, WEE1 and MCL-1) at the lowest dose level (7.5 mg) (**Figure 3b**).

**Figure 4.**
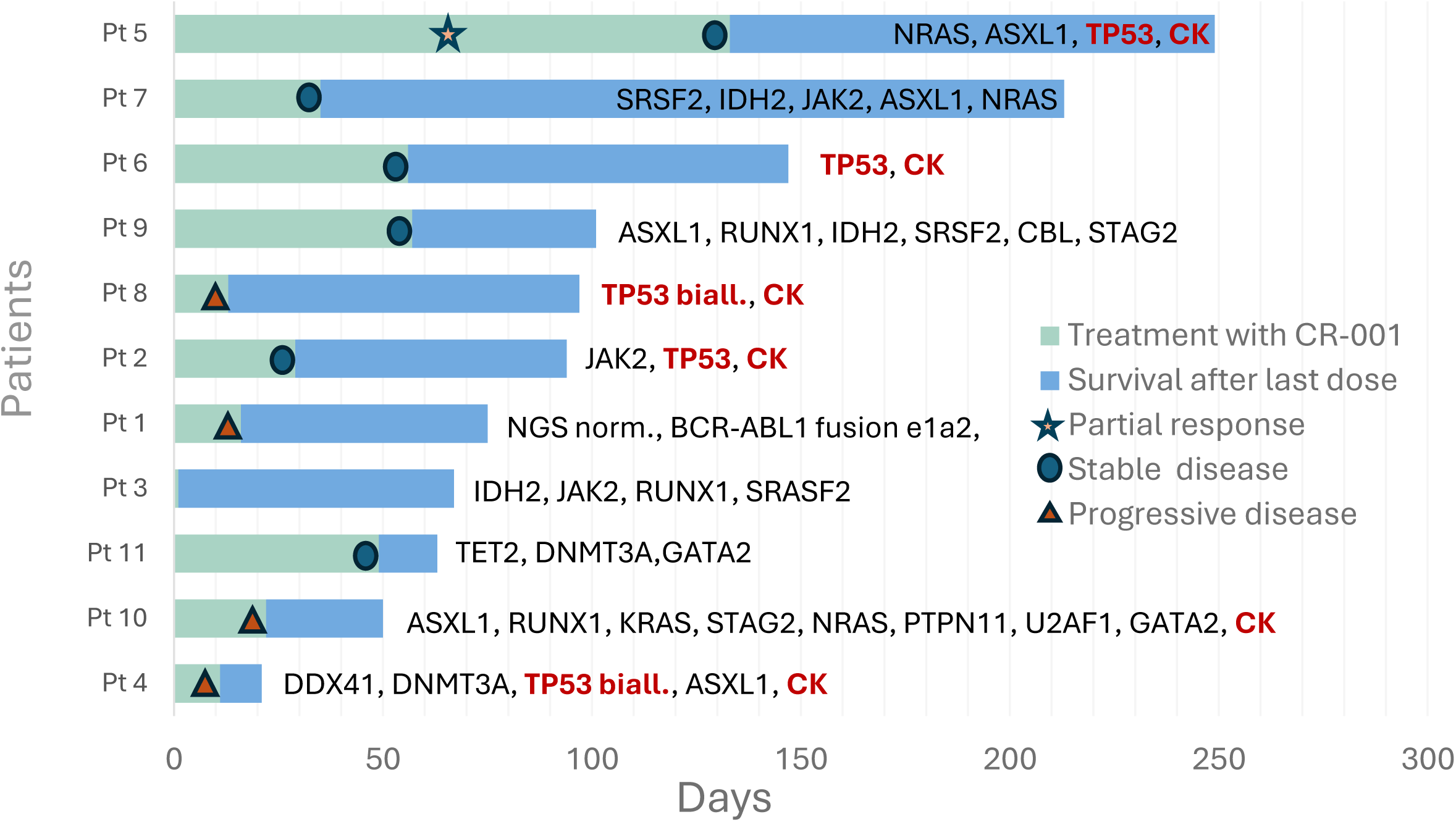
Swimmer plot of study participants. Pt 1 – Pt 4 received dose level 1 (7.5 mg). Pt 5 – Pt 7 received dose level 2 (11.25 mg). Pt 8 – Pt 10 received dose level 3 (15 mg). Pt 11 received dose level 4 (18.75 mg).

**Figure 3b.**
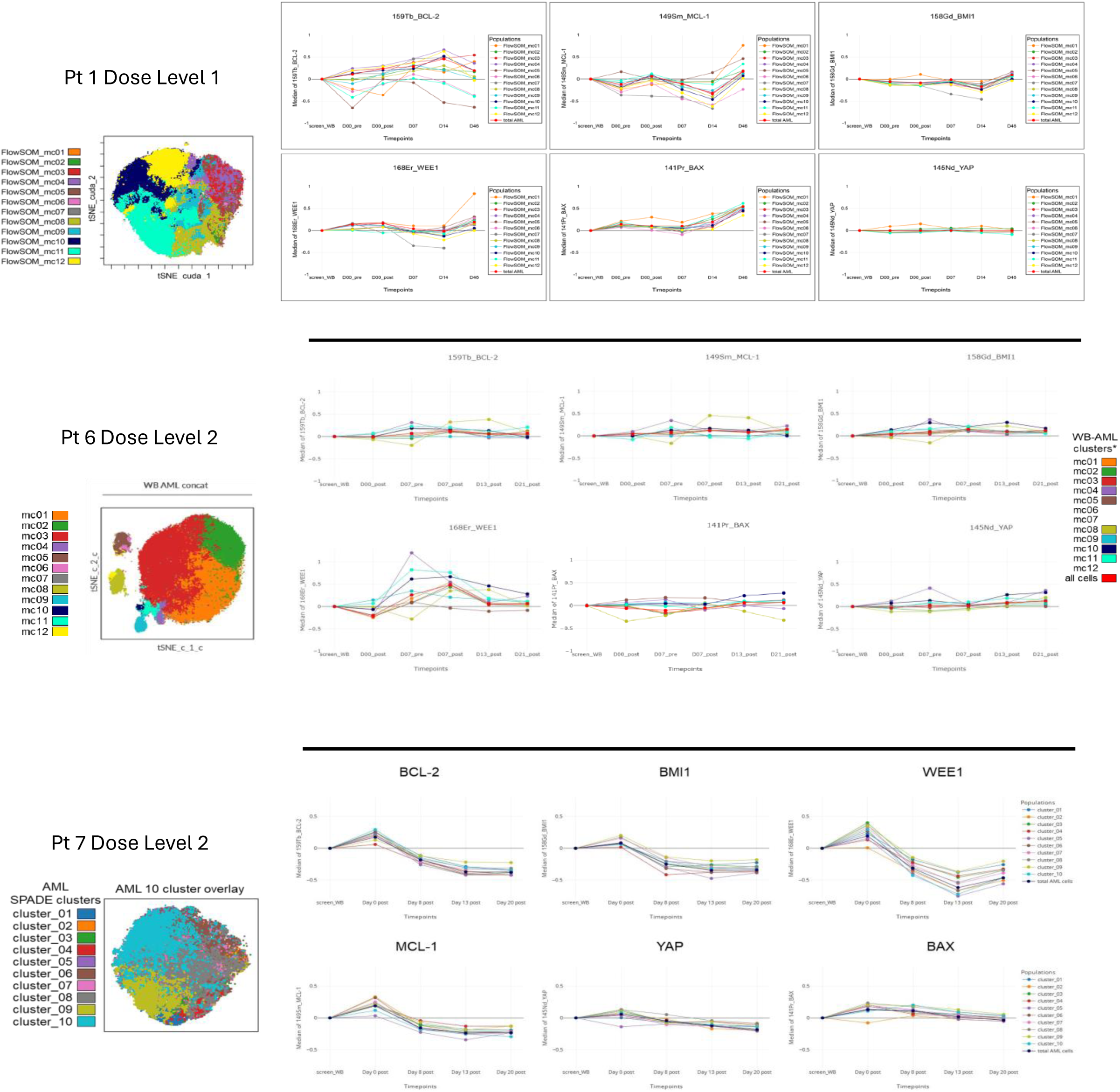
Median expression of key proteins in whole blood (WB) AML clusters. AML cells were identified using mass cytometry. The cells were clustered per patient using FlowSOM or SPADE. The median expression of each protein in every cluster of every patient was calculated for every timepoint relative to screening timepoint. *Clusters with fewer than 150 cells in every sample were excluded from this figure.

Preliminary efficacy was evaluable in 10 (91%) patients, all receiving at least three doses of CR-001. Six (60%) evaluable patients achieved stable disease (ELN 2022) after 3-4 doses (patients #2, 5, 6, 7, 9 and 11), and two patients remained with stable disease after 8 doses (patients #5 and 6). Bone marrow aspirate count reduction after 3-4 infusions was observed in 3 (30%) patients after 3-4 doses (patients #6, 7 and 9), and in one (10%) patient after 8 doses (patient #5).

Progressive disease was noted in 4 (40%) patients after 3-4 doses. The patient with breast cancer therapy-related AML with complex karyotype, *TP53* mutation, and relapse after a second allogeneic stem cell transplant, experienced lasting regression of leukemic pericardial effusion after two doses, excellent performance status, and survived for 5.5 months after the last (16^th^) dosage of CR-001 at dose level 11.25 mg. Signs of granulopoiesis maturation/differentiation were observed in the bone marrow both morphologically on smear microscopy and immunophenotypic flowcytometry. This was correlated with increase/normalization of neutrophils in the blood. The patient received concomitant hydroxyurea during this study and entered an experimental trametinib treatment after disease progression (*NRAS* mutation). Increase/normalization of neutrophils was observed in one more patient at the same dose level who also achieved stable disease after 4 doses.

### Correlative endpoints

The mass cytometry results demonstrated that the expression of WEE1, BCL2, MCL1 were slightly reduced by the treatment with CR-001 at the lowest dose level (7.5 mg) (**Figure 3b**). At the same time BAX, a marker of pro-apoptosis, increased following treatment. While a more pronounced reduction in WEE1, BCL2, and MCL1 was observed at a higher dose level (11.25 mg) in patient 7 , another patient 6 at the same dose level (11.25 mg) did not reproduce the same effect.

## Discussion

This multicenter, first-in-human, phase I clinical trial is the first to investigate safety, tolerability, and preliminary efficacy of CR-001, a novel 5-fluorouracil (5-FU)–modified miR-15a mimic, in R/R AML patients.

Treatment of 11 AML patients with CR-001 was well-tolerated across the 4 dose levels administered on an out-patient basis. All treated patients had an adverse event (AE) during treatment. We observed no dose-limiting toxicities. The most frequent AE was infection and was reported in all patients. The second most frequent AE was transient hyperglycemia, been attributed to the use of oral steroids as premedication prior to infusion. 6 patients experienced infusion-related reaction experiencing chills and tachycardia lasting shortly and resolving completely upon intravenous administration of hydrocortisone and antihistamine intravenously. This reaction subsided in most patients after the infusion time was increased to 2 hours. Cytokine release syndrome (CRS) was suspected in one patient due to fever as an additional symptom, and received therefore tocilizumab, an interleukin blocker. Performance status was improved in 3 patients and in total 8 patients had an ECOG score 0-1 after 4 doses. Preliminary clinical activity of CR-001 was observed in 6 of 10 evaluable patients, who achieved ELN 2022-defined stable disease after 3–4 doses; one maintained stability after 8 doses. Disease control was most notable in patients with *TP53* mutations and complex karyotypes. One patient with therapy-related AML and *TP53* mutation showed elimination of pericardial AML after four doses and received in total 16 doses, surviving 5.5 months post-treatment. Signs of granulocytic maturation and normalization of neutrophils were noted in this case and one additional patient at the same dose level.

Single-cell mass cytometry confirmed on-target activity of CR-001, including downregulation of miR-15a targets (BMI1, WEE1, MCL-1) and increased pro-apoptotic BAX expression at the lowest dose level (7.5 mg**) (Figure 3b)**. These early data suggest that CR-001 may exert disease-stabilizing and pro-differentiation effects, particularly in genomically defined high-risk AML. ^18^

Several scientific reports provide insights into the role of miR-15a in AML and the development of 5-FU-modified miRNAs as cancer therapeutics. ^10–14^ It has been reported that knockout of both miR-15/16-1 and miR-15b/16-2 clusters drives the development of aggressive AML. ^11^ Beyond AML, we have also demonstrated that restoration of miR-15a exerts antitumor activity in several solid malignancies, including colorectal and pancreatic cancer, through suppression of multiple oncogenic targets ^12,14^. These targets include BCL2, YAP1, WEE1, CHK1, and BMI1-several of which play known roles in chemoresistance and leukemic cell survival. Notably, BCL2 is a well-established miR-15a target in both CLL and AML . ^11,12^ YAP1 has been implicated in doxorubicin resistance in AML. ^21^ Combined suppression of CHK1 and WEE1 has demonstrated synergistic antileukemic effects in AML *ex vivo*. ^22^ Moreover, inhibition of BMI1 has been shown to induce p53-independent mitochondrial apoptosis in AML progenitor cells. ^23^ CR-001 was well-tolerated by R/R AML patients (**Table 2)**. Such results establish a potential for a safe and well-tolerated treatment regimen for AML patients who may choose to stay for treatment with more treatment cycles to maximize therapeutic benefits.

Based on these reports, we hypothesized that the multi-targeted CR-001 could provide a unique therapeutic advantage in suppressing pathways driving AML resistance and relapse. Single-cell profiling using mass cytometry in this trial supports this hypothesis: molecular activity of CR-001 was detected on canonical miR-15a targets (e.g., BMI1, WEE1, MCL-1) even at the lowest dose level (7.5 mg) (**Figure 3b**).

To develop a clinically viable miR-15a-based therapeutic strategy, we have previously developed a tumor suppressor miRNA-based platform technology in which substitution with 5-FU enhances both therapeutic potency and intracellular delivery in various solid tumor types and AML. ^14^ The addition of 5-FU increases molecular lipophilicity, an extensively applied method in small molecule drug development, and improves cell membrane permeability of otherwise negatively charged RNA molecules. The 5-FU-modification thus not only retains anticancer activity but also facilitates cellular uptake of the miRNA mimic, contributing to a dual mechanism of action. Our strategy takes advantage of 5-FU as an active anti-cancer therapeutic compound and the fluorine group also enhances the deliverability of a nucleic-acid based miRNA tumor suppressor. ^14^ Once CR-001 (5-FU-miR-15a) breaks down by 5’-and 3’-exonucleases, it will release chemotherapeutic agent 5-FU to exert its therapeutic activity by suppressing its target protein thymidylate synthase. 5-FU modification of miRNAs offers unique advantages as cancer therapeutics compared to single target agents by suppressing multiple oncogenic targets and pathways. This will be a potential superior feature to overcoming drug resistance, offering major survival benefits and less chance of disease relapse. ^9^

The CR-001 formulation includes a small amount of polyethyleneimine (PEI) as a delivery vehicle formulation to further enhance the therapeutic efficacy of miR-15a in AML.

The rationale to include PEI was based on previous findings that PEI can further enhance the potency of CR-001 in colorectal cancer treatment. ^14^ As PEI can be potentially toxic if given at higher doses, we designed the Phase I study strategically by escalating the dose of CR-001 while keeping the dose of the delivery vehicle, PEI, constant at a low level.

This approach ensures that we can maximize the therapeutic efficacy of CR-001 and at the same time preserve the delivery advantages of PEI without introducing the potential toxic effect of PEI as the vehicle. It is clearly demonstrated that our phase I AML patients have well tolerated the dose escalation of CR-001.

Overall, CR-001 demonstrated a favorable safety profile, with no dose-limiting toxicities (DLTs) observed across all dose levels. Notably, no grade 4 treatment-related adverse events were reported, and grade 3 events were infrequent, non-cumulative, and manageable with standard supportive care measures. Importantly, no patients discontinued treatment due to adverse events, and there were no treatment-related deaths. The absence of severe organ dysfunction is particularly encouraging, suggesting that the compound may be well tolerated even in clinically vulnerable populations. High-grade cytopenias and deaths were not related to the treatment but to the disease progression of the patients who did not respond to the investigational drug. These findings suggest that the investigational agent may offer a safer alternative to existing therapies, particularly for vulnerable patient populations such as the elderly or those with comorbidities, who often cannot tolerate standard cytotoxic regimens.

In summary, CR1-02 is the first-in-human phase I clinical trial based on tumor suppressor miR-15a in AML. CR-001 was well-tolerated and showed signals of biological activity as evidenced by disease stabilization when administered in relapsed or refractory AML patients. There are signs of biological activity beginning at the lowest dose given (7.5 mg/administration), and clinical efficacy signals starting at 11.25 mg/administration. Based on its superior safety profile, CR-001 may have a potential as an ideal candidate for AML maintenance therapy. Future development activities will focus on combination with other therapeutic agents to maximize the benefit to AML patients.

## Data Availability

All data produced in the present study are available upon reasonable request to the authors.

## Acknowledgments

We would like to express our deepest gratitude to the patients enrolled in this study. The study was funded by Curamir Therapeutics Inc., as well as Smerud Medical Research International AS, KinN Therapeutics AS, Haukeland University Hospital and Oslo University Hospital.

---

**Supplementary table 1:** 39-marker mass cytometry antibody panel. IC: Intracellular

| **Target** | **Clone** | **Mass tag** | **Staining layer** | **Cat. #** | **Vendor** |
| --- | --- | --- | --- | --- | --- |
| CD45 | HI30 | 89Y | surface | 3089003B | Standard Bitools |
| CD3 | UCHT1 | 111Cd | surface | 300443 | BioLegend |
| CD8 | HIT8a | 113Cd | surface | 300902 | BioLegend |
| CD19 | HIB19 | 142Nd | surface | 3142001B | Standard Bitools |
| CD38 | HIT2 | 144Nd | surface | 3144014B | Standard Bitools |
| CD64 | 10.1 | 146Nd | surface | 3146006B | Standard Bitools |
| CD13 | WM15 | 147Sm | surface | 3147014B | Standard Bitools |
| CD274/PD-L1 | 29E.2A3 | 148Nd | surface | 3148017B | Standard Bitools |
| CD123/IL-3R | 6H6 | 151Eu | surface | 3151001B | Standard Bitools |
| CD36 | 5-271 | 152Sm | surface | 3152007B | Standard Bitools |
| CD7 | CD7-6B7 | 153Eu | surface | 3153014B | Standard Bitools |
| CD117/c-kit | 104D2 | 154Sm | surface | 313223 | BioLegend |
| CD56/NCAM | B159 | 155Gd | surface | 3155008B | Standard Bitools |
| CD14 | RMO52 | 160Gd | surface | 3160006B | Standard Bitools |
| CD90/Thy-1 | 5E10 | 161Dy | surface | 3161009B | Standard Bitools |
| CD11c | Bu15 | 162Dy | surface | 3162005B | Standard Bitools |
| CD33 | WM53 | 163Dy | surface | 3163023B | Standard Bitools |
| CD15/SSEA-1 | W6D3 | 164Dy | surface | 3164001B | Standard Bitools |
| CD16 | B73.1 | 165Ho | surface | 3165007B | Standard Bitools |
| CD34 | 581 | 166Er | surface | 3166012B | Standard Bitools |
| CD25/IL-2R | 2A3 | 169Tm | surface | 3169003B | Standard Bitools |
| CD45RA | HI100 | 170Er | surface | 3170010B | Standard Bitools |
| CD44 | IM7 | 171Yb | surface | 3171003B | Standard Bitools |
| CD371/CLEC12A | 50C1 | 173Yb | surface | 3173007B | Standard Bitools |
| HLA-DR | L243 | 174Yb | surface | 3174001B | Standard Biotoools |
| CD279/PD-1 | EH12.2H7 | 175Lu | surface | 3175008B | Standard Biotoools |
| CD4 | RPA-T4 | 176Yb | surface | 3176010B | Standard Biotoools |
| CD11b/Mac-1 | ICRF44 | 209Bi | surface | 3209003B | Standard Biotoools |
| BAX | 2D2 | 141Pr | IC | 633602 | BioLegend |
| c-PARP | F21-852 | 143Nd | IC | 3143011A | Standard Biotoools |
| YAP | 1A12 | 145Nd | IC | NBP2-22117 | Novus Biologicals |
| MCL-1 | D2W9E | 149Sm | IC | 94296BF | Cell Signaling Technologies |
| p53 | DO7 | 150Nd | IC | 3150024A | Standard Biotoools |
| p-p38 [T180/Y182] | D3F9 | 156Gd | IC | 3156002A | Standard Biotoools |
| BMI1 | D20B7 | 158Gd | IC | 6964BF | Cell Signaling Technologies |
| BCL-2 | 124 | 159Tb | IC | NBP2-34513-0.1mg | Novus Biologicals |
| p-ERK1/2 [T202/Y204] | D13.14.4E | 167Er | IC | 3167005A | Standard Biotoools |
| WEE1 | D10D2 | 168Er | IC | 13084BF | Cell Signaling Technologies |
| Ki-67 | B56 | 172Yb | IC | 3172024B | Standard Biotoools |

